# Estimation of the morbidity and mortality of congenital Chagas disease: a systematic review and meta-analysis

**DOI:** 10.1101/2022.04.12.22273277

**Authors:** Sarah Matthews, Ayzsa F. Tannis, Karl Philipp Puchner, Maria Elena Bottazzi, Maria Luisa Cafferata, Daniel Comandé, Pierre Buekens

## Abstract

Chagas disease is caused by the parasite *Trypanosoma cruzi* which can be transmitted from mother to baby during pregnancy. There is no consensus on the proportion of infected infants that become symptomatic for congenital Chagas disease (cCD). The objective of this systematic review is to determine the burden of cCD.

Articles from journal inception to 2020 reporting morbidity and mortality associated with cCD were retrieved from academic search databases. Observational studies, randomized-control trials, and studies of babies diagnosed with cCD were included. Studies were excluded if they were case reports or series, without original data, case-control without cCD incidence estimates, and/or did not report number of participants. Two reviewers screened articles for inclusion. To determine pooled proportion of symptomatic infants with cCD, individual symptoms, and case-fatality, random effects meta-analysis was performed.

We identified 4,531 records and reviewed 4,301, including 47 articles in the narrative summary and analysis. 28.3% (95% confidence interval (CI) = 19.0%, 38.5%); of cCD infants were symptomatic and 2.2% of infants died (95% CI = 1.3%, 3.5%). The proportion of infected infants with hepatosplenomegaly was 12.5%, preterm birth 6.0%, low birth weight 5.8%, anemia 4.9%, and jaundice 4.7%. Although most studies did not include a comparison group of non-infected infants, the proportion of infants with cCD symptomatic at birth are comparable to those with congenital toxoplasmosis (10.0%-30.0%) and congenital cytomegalovirus (10.0%-15.0%).

We conclude that cCD burden appears significant, but more studies comparing infected mother-infant dyads to non-infected ones are needed to determine an association of this burden to cCD infection.

**Author summary:** Chagas disease is caused by the parasite *Trypanosoma cruzi*, which can be passed from mother to infant. It is estimated that one million women of reproductive age are infected with *T. cruzi*. Prior to our work, the proportion of infants infected with *T. cruzi* congenitally presenting with clinical symptoms was unknown. After systematically searching for and identifying studies that collected information on infants with congenital Chagas disease, we summarized and analyzed 47 studies. Our pooled analysis of these studies estimated that 28.3% of infants with congenital Chagas disease were symptomatic and 2.2% died. Prior work has shown that transmission of *T. cruzi* from mother to child occurs in 5% of cases. Other studies have shown that this transmission is preventable through treatment of women prior to conception, and infants can be cured if shown to be infected at birth. Our estimated proportion of 28.3% of infants diagnosed with cCD at birth presenting with clinical symptoms are comparable to infants diagnosed with congenital toxoplasmosis presenting with clinical symptoms (10.0%-30.0%) and congenital cytomegalovirus (10.0%-15.0%). More studies comparing infected mother-infant dyads to non-infected mother-infant dyads are needed to determine an association of this burden to cCD infection.

## Introduction

### Background

Chagas disease, caused by the protozoan parasite *Trypanosoma cruzi*, is estimated to infect 6.5 million globally, including 1.7 million women of reproductive age [1, 2]. As of 2019, an estimated 172,000 additional people were infected, and 52,000 of these were women of reproductive age [1]. *T. cruzi* is primarily transmitted when the triatomine insect vector transfers the parasite after biting and defecating on its host through its infected feces entering via bite wound or mucosal membrane [2]. However, it can also be transmitted through blood transfusion, organ transplant, via oral consumption of contaminated food with triatomine feces, and through vertical transmission from mother to infant during pregnancy [3, 4].

Vertical transmission of *T. cruzi*, or congenital Chagas disease (cCD) occurs in an estimated 4.7% of infants born to infected mothers, increasing to 5.0% in endemic countries [3]. *T. cruzi* infected infants may present with severe morbidity at birth and be at a higher risk of mortality. If left untreated, infants can develop chronic Chagas disease later in life [4].

### Signs and symptoms of congenital Chagas disease

Signs and symptoms in cCD infected infants range from mild to severe. Signs attributable to cCD include low Apgar score (<7 at 1 minute and/or at 5 minutes) [5], premature rupture of membranes [6], preterm birth, and low birth weight [7], intra-uterine growth restriction [8], small for gestational age [9], and neonatal intensive care unit (ICU) admission [10]. Mortality attributed to cCD is associated with severe morbidity symptoms, including meningoencephalitis and myocarditis [2]. Reported symptoms attributable to cCD are hepatomegaly, splenomegaly, respiratory distress syndrome, certain neurologic signs, anasarca, petechiae, abnormal electrocardiographic findings, anemia, meningoencephalitis, myocarditis, congestive heart failure, digestive and/or central nervous system lesions, parasites in various tissues [2], subependymal hemorrhage [11], and cardiomegaly [6].

### Clinical pathway

Infants exposed in-utero to *T. cruzi* are susceptible to congenital transmission [12]. Screening programs to diagnose and treat pregnant women and infants infected with *T. cruzi* have been implemented in endemic countries and countries with large migrant populations from endemic regions since the early 1990s [13, 14]. The gold standard for diagnosing acute and chronic infection uses at least two conventional serological tests (e.g., indirect hemagglutination assay, indirect immunofluorescence assay, ELISA) [12, 15]. Other tests, such as molecular tests and rapid diagnostic tests can also confirm infection but are only recommended to complement or confirm aforementioned assays [12, 15].

Confirmation of cCD in infants born to *T. cruzi* infected mothers occurs at birth or in the first weeks afterward by viewing parasites in an umbilical cord blood sample or venous infant blood, or after 8-10 months when maternal antibodies have waned using serological assays to confirm infant *T. cruzi* IgG antibodies [4, 12]. Gold standard diagnosis of cCD requires at birth, parasitological examination using microhematocrit or microstrout testing methods, and if negative repeated examination one month later, and at 10 months two serological tests [12]. Molecular methods of diagnosis can detect infection early on but are not part of the gold standard diagnosis given lack of standardization, low and often fluctuating parasitemia in patients with chronic Chagas disease and lack of quality control programs [16].

Evidence shows that if women are treated for Chagas disease before pregnancy, future congenital transmission of *T. cruzi* is preventable [4]. Treatment during pregnancy is not recommended given unknown effects of antiparasitic drugs on prenatal development. Treatment of *T. cruzi* infected infants with benznidazole or nifurtimox is effective when administered within the first year of life [17].

#### Rationale

In 2010, its estimated that between 158,000 to 214,000 infants were born to *T. cruzi* infected mothers in endemic countries, of which 8,000 to 10,700 would be congenitally infected [18]. Around 1/5 of annual new Chagas cases are attributed to congenital infection [16]. The Global Burden of Disease project used data from vital registration databases, surveillance, surveys/census, and other population-based sources to estimate the burden of Chagas disease among neonates and infants, including number of deaths, disability-adjusted life years, years lived with disability, and years of life lost [19]. However, it is likely that the data used to estimate the burden of Chagas disease are incomplete given issues in diagnosing cCD, including low sensitivity of parasitological screening at birth and loss to follow-up with serological screening 8-10 months postpartum [18]. Given this, there is no accurate burden estimate for cCD and no consensus on how many infants are symptomatic [2]. The objective of this systematic review is to determine the morbidity and mortality of cCD.

## Methods

A systematic review and meta-analysis were performed according to the guidelines of the Meta-Analysis of Observational Studies in Epidemiology (MOOSE) and the Preferred Reporting Items for Systematic reviews and Meta-Analysis (PRISMA) [20, 21]. The protocol was registered on PROSPERO [22].

### Criteria for considering studies

#### Types of studies

Studies that reported morbidity or mortality associated with cCD infection were considered, including observational studies and randomized-control trials. Studies excluded were case reports and series, studies not including original data were excluded, case-control studies without neonatal incidence estimates of cCD, and studies not reporting the number of infected neonates.

#### Types of participants

Studies about diagnosed neonates and infants with cCD were included.

#### Types of outcomes

Articles including original data of morbidity or mortality among infants with cCD were included. Mortality was defined as the recorded death of a *T. cruzi* infected fetus or infant. Morbidity was defined as any adverse outcome presenting in a *T. cruzi* infected infant, with all symptoms extracted available in the **S1 File**. Mortality causes included stillbirth, miscarriage, abortion, intrauterine death, and fetal death.

### Search strategy

A medical librarian developed and applied a comprehensive and sensitive search strategy (available in **S2 File**) using terms related to cCD in PubMed, EMBASE, CINAHL, LILACS, and Academic Search databases. No language restrictions were applied, and grey literature was not searched.

### Data collection and analysis

#### Selection of studies

Authors AT and SM independently screened study titles and abstracts and then the remaining full text articles for eligibility. All disagreements were resolved by discussion and, if necessary, a third author (KP) was consulted as an arbitrator. Covidence systematic review software was used to facilitate the screening process [23]. For duplicate studies, the one with the largest sample size was included. The decision-making algorithm consideration is available in **S3 File**.

#### Data extraction and management

Authors AT and SM independently extracted data using a form designed and piloted with studies a priori. Extracted data included study, maternal, and infant characteristics, diagnostic information for mothers and infants, and morbidity and mortality of congenital cases. A summary of extracted data can be found in **S4 File** and the data extraction form in **S1 Dataset**.

Data extraction discrepancies were resolved by discussion and, if necessary, a third author (KP) was consulted. The inter-observer reviewer agreement for full text screening was assessed using the Kappa statistic.

#### Assessment of risk of bias

A risk of bias assessment tool was developed through adaptation of the NIH Study Quality Assessment Tools and the STROBE (Strengthening the Reporting of Observational studies in Epidemiology) checklist of essential items for observational studies [24, 25]. Authors AT and SM piloted the tool on five studies, subsequently adapted the tool and then independently assessed included studies’ risk of bias of the included studies (**S5 File)**. Six domains were considered: 1) participant selection methods, 2) exposure and outcome variable measurement, 3) confounding control methods, 4) reporting of results, 5) statistical methods, and 6) declaration of conflict and ethical statements. Two algorithms were developed to summarize within-domain and summary risk of bias (**S5 File)**.

#### Statistical analysis and data synthesis

Included study frequencies of congenital transmission, symptoms, mortality causes (including those not originally listed in **S1 File**), infant mortality and/or case-fatality rates, and proportion of asymptomatic and symptomatic cCD was narratively summarized.

A meta-analysis of proportions was performed to estimate the pooled proportion of symptomatic fetuses and infants with cCD. The Freeman-Tukey double arcsine method was used to account for overdispersion of proportions and stabilize the variance [26, 27]. Stuart-Ord inverse variance weight were applied to transformed proportions, avoiding underestimation of true variance using its conservative weight [28]. The pooled proportion and its 95% confidence interval (CI) were estimated using the DerSimonian-Laird random effects model to take into consideration the high likelihood of between-study heterogeneity. Results were quantified and represented in a forest plot [28, 29]. The proportion of symptomatic cCD cases was defined as the number of infants with cCD displaying symptom(s) and/or death divided by the total number of infants with cCD. We also performed a meta-analysis of proportions for the pooled proportion of death due to cCD. If a study reported cCD symptoms and/or mortality frequency but did not provide a frequency for every outcome outlined in **S1 File** and/or death, missing values were assumed to be non-events and a value of 0 was imputed [30]. All analyses were performed using SAS Version 9.4, Stats-Direct, and StataIC 12 software.

#### Assessment of heterogeneity

The I^2^ statistic was calculated to measure the proportion of total variability attributable to heterogeneity between studies [31]. Three subgroup analyses defined a priori were performed by: cCD diagnostic method, geographic region, and individual symptom displayed in fetus/infant. Studies were excluded for the subgroup analysis of symptom frequency if no clear definition of each symptom displayed in individual infants was reported. A subgroup of co-infection with other non-Chagas related infections was planned; however, data was insufficient. Detailed results are described in **S6 File**.

#### Sensitivity analyses

Two sensitivity analyses were conducted to assess the potential effect review decisions held on robustness of results. These analyses were to exclude studies with high risk of bias and exclude studies where Chagas disease gold standard diagnosis of the mother was not employed (15). An ad-hoc sensitivity analysis was performed using the Miller back-transformation [32] for the primary meta-analyses of proportion of cCD morbidity and mortality. Detailed results are described in **S6 File**.

#### Assessment of publication bias

The effect of publication bias was evaluated for all analyses using Egger’s statistical test to determine asymmetry of the funnel plot [33].

## Results

A total of 4,531 records were identified through database search, 4,301 were screened based on title and abstract, and 293 full text articles assessed for eligibility, and 47 articles were included for narrative summary and meta-analysis.

### Narrative summary

Study publication year ranged from 1962 to 2019, with 18 studies published before 2000, 10 between 2000 to 2010, and 18 between 2011 and 2019. Study duration ranged from under one year to 15 years, with six studies under one year, 31 studies one to four years, and eight studies five to 15 years, with two studies missing data on this factor. 12 studies were conducted in Europe and 35 in the Latin American and Caribbean region (Mexico, Central and South America). Most studies (n=38) were conducted in urban/semi-urban hospital(s), with one conducted in a rural hospital, four in both rural and urban hospitals, one conducted in primary care institutions, and three with missing information. In regard to study design, three were case-control, 13 cross-sectional, 28 prospective cohorts, two retrospective cohorts, and one a mixed cohort. Study population size varied from eight to 4,355 infants. 15 studies had less than 100 infants, 14 had between 100 and 999 infants, and 10 had over 1,000 infants, with five studies missing data. 28 studies used gold standard diagnosis for mothers, 16 used an alternative, and three studies did not provide information. 13 studies diagnosed infants with cCD using the gold standard, 32 used an alternative, and two did not provide information. **Table 1** summarizes all study characteristics.

**Table 1:**
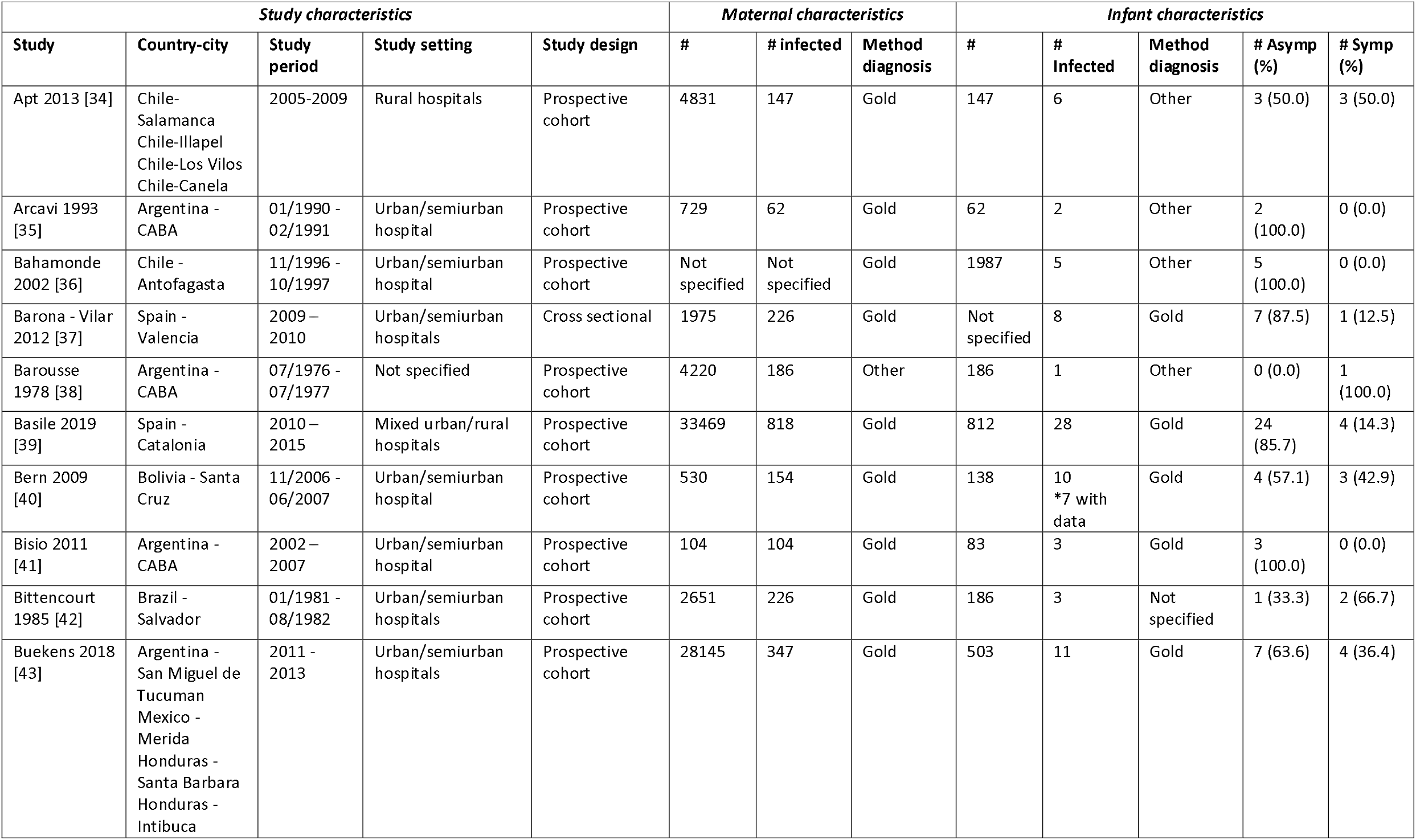
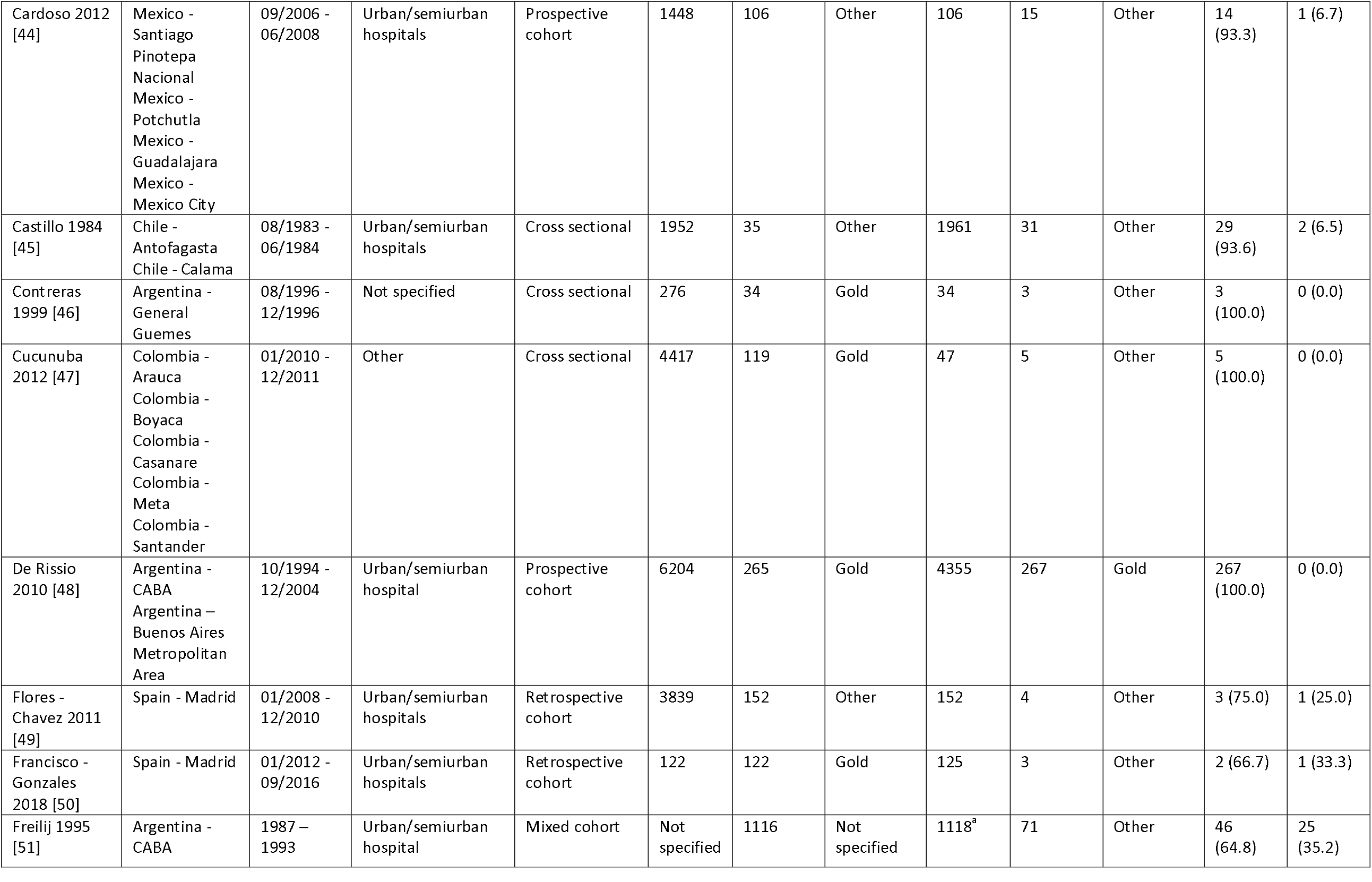
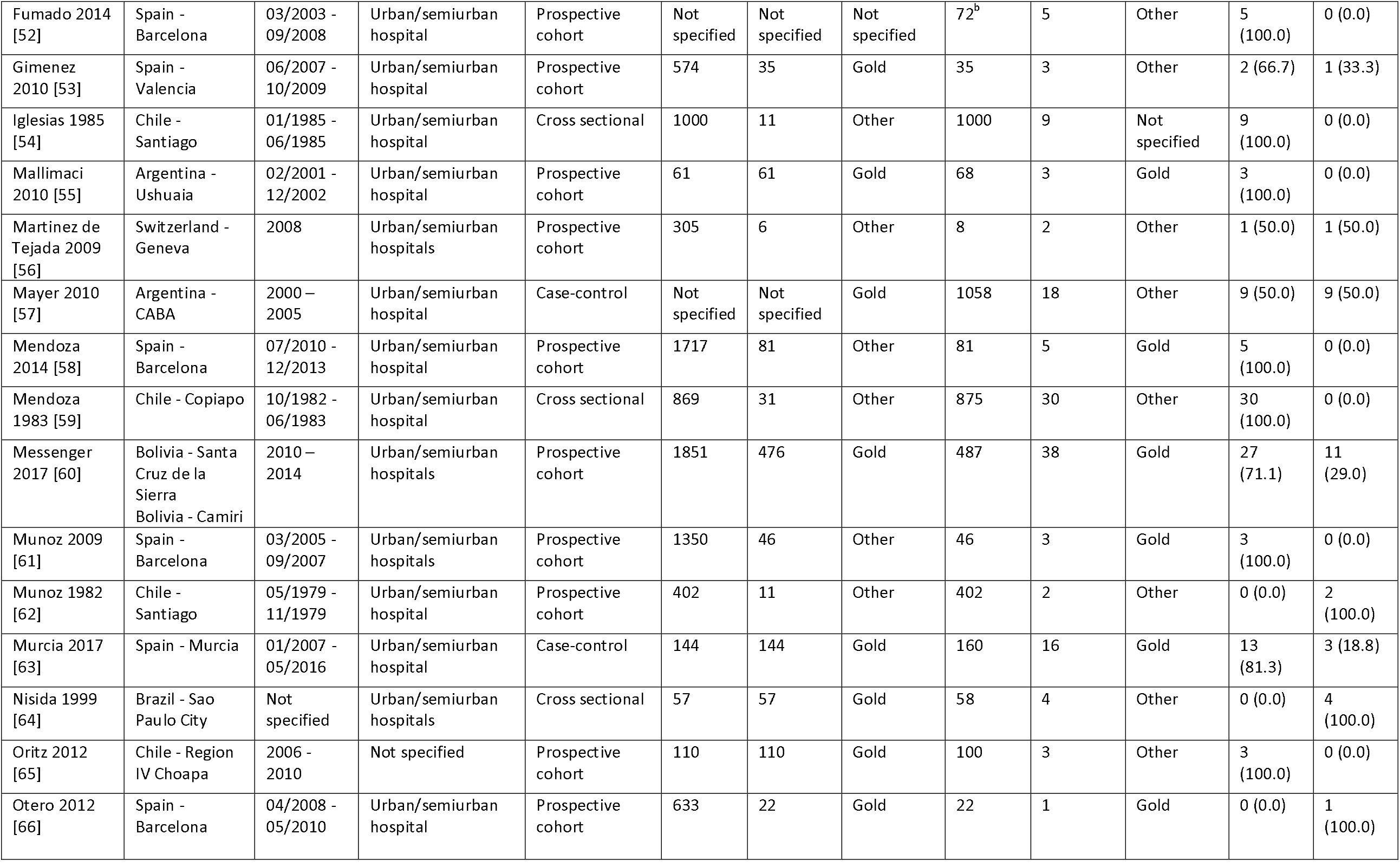
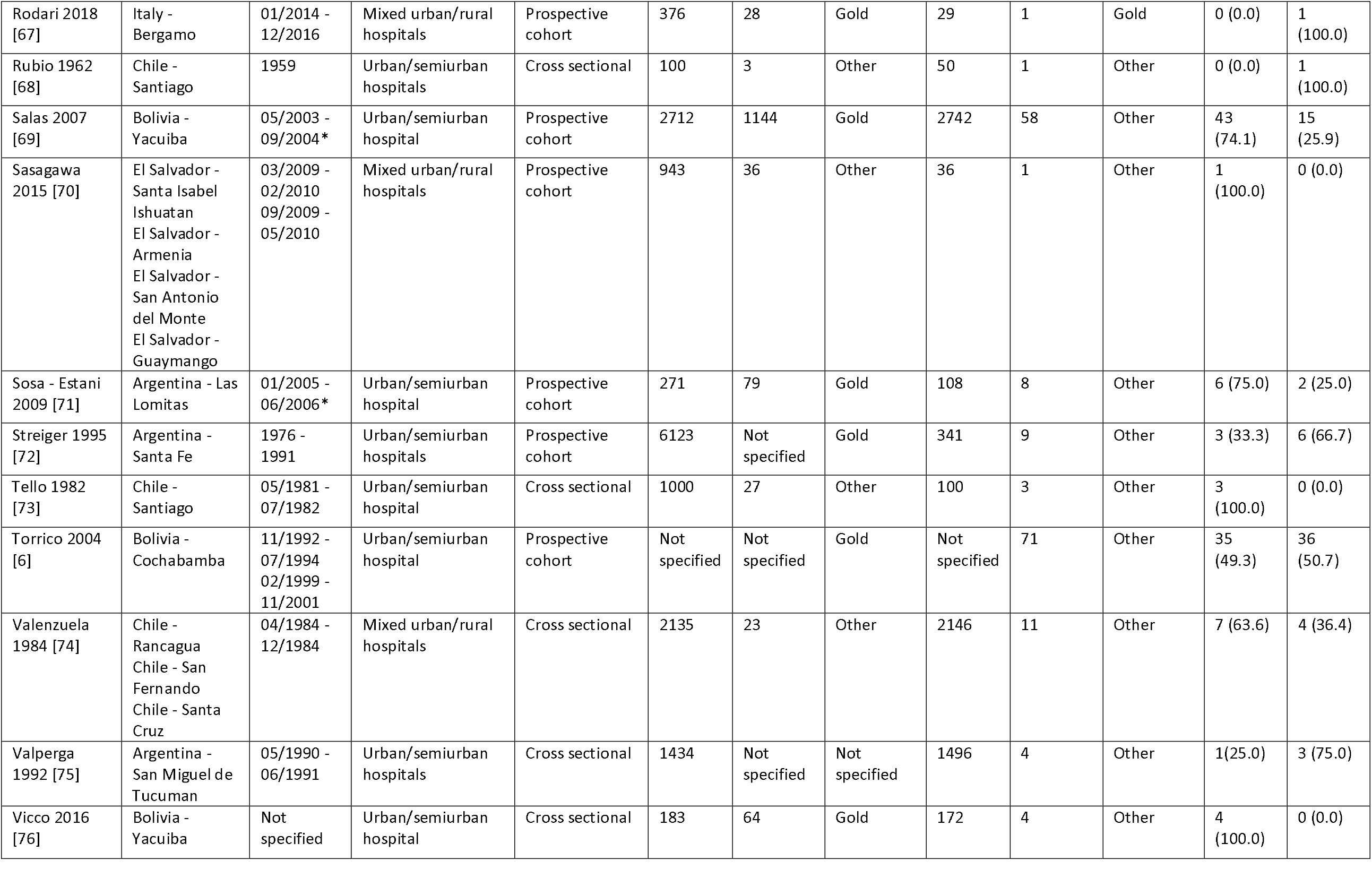
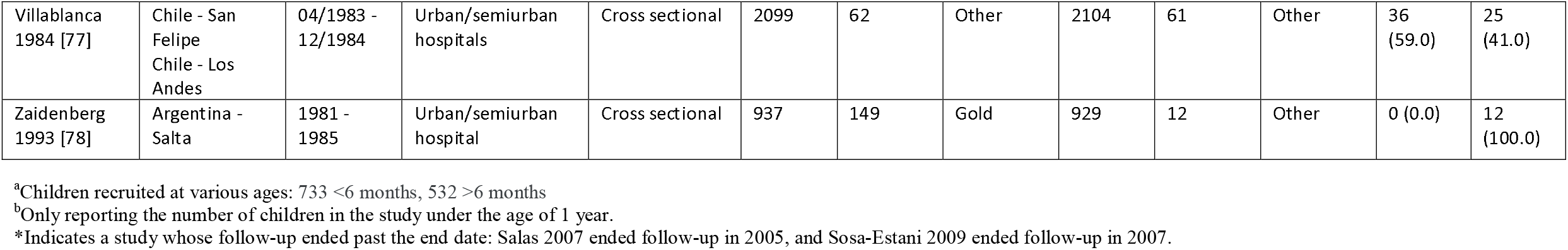
Study Characteristics.

The number of cCD infected infants in studies ranged from one to 267, with a median of five. There were 25 studies with five or less cCD infected infants, six with six to 10 infected infants, 11 with 11 to 50 infected infants, four with 50 to 100 infected infants, and one with over 100 infected infants. The percentage of symptomatic infected infants among all infected infants ranged from 0.0% to 100.0%, with a median of 26.0%. 16 studies reported a percentage of 0.0%, five reported 1.0% to 25.0%, 12 reported 26.0% to 50.0%, four reported 51.0% to 99.0%, and seven reported 100.0% symptomatic. Symptoms of cCD by study, including their reported frequency, are in **S1 Table**. Eight studies reported infant mortality for cCD, three citing Chagas as cause of death, with other causes being stillbirth, Down’s syndrome, congenital cardiopathy, respiratory distress, severe neurological damage, gastroenteritis and dehydration, pneumonia, and secondary septicaemia and pneumococcal meningitis. One study reported four deaths, four studies each reported two infant deaths, and the remaining three reported one infant death. Time of death ranged from birth to 14 months. cCD mortality characteristics are in **S2 Table**.

### Primary analyses

Results from the primary analyses can be found in **Table 2**. The primary meta-analysis of the proportion of symptomatic cCD infected infants to all cCD infected infants revealed a pooled proportion of 28.3% (95% CI = 19.0%, 38.5%). This estimate had a I^2^ inconsistency statistic of 88.6% (95% CI = 86.0%, 90.5%), suggesting considerable heterogeneity between studies for morbidity. The Egger’s bias statistic was statistically significant (P < 0.0001), suggesting that publication bias influenced these results. The forest plot and Egger’s bias plot can be viewed in **Figs 1 and 2**.

**Table 2.** Primary Analyses and Subgroup Analyses.

| | Pooled<br>Proportion<br>% | 95% CI | $I^2$ (%) | 95% CI | Egger's<br>Bias | P-Value* |
| --- | --- | --- | --- | --- | --- | --- |
| **Primary Analyses (N=47)** |  |  |  |  |  |  |
| Morbidity | 28.3 | 19.0,38.5 | 88.6 | 86.0,90.5 | 2.5 | <0.0001 |
| Mortality | 2.2 | 1.3,3.5 | 9.6 | 0.0,37.5 | 0.3 | 0.01 |
| **Subgroup 1 (N=45)** |  |  |  |  |  |  |
| Infant gold (n=13) | 18.7 | 6.1,36.1 | 86.8 | 79.2,90.8 | 1.8 | 0.00 |
| Infant other (n=32) | 32.5 | 23.0,42.9 | 78.9 | 70.5,84.1 | 1.8 | 0.08 |
| **Subgroup 2 (N=47)** |  |  |  |  |  |  |
| Europe (n=12) | 20.0 | 11.4,30.2 | 10.2 | 0.0,54.8 | 2.3 | 0.04 |
| Latin America and the Caribbean (n=35) | 29.4 | 18.3,41.8 | 91.3 | 89.3,92.7 | 2.9 | <0.0001 |
| **Subgroup 3 (N=47)** |  |  |  |  |  |  |
| Hepatosplenomegaly | 12.5 | 6.6,19.9 | 85.8 | 82.2,88.4 | 1.4 | 0.003 |
| Preterm birth | 6.0 | 3.3,9.5 | 61.2 | 44.5,71.1 | 0.8 | 0.0003 |
| Low birth weight | 5.8 | 3.2,9.1 | 60.7 | 43.8,70.8 | 0.7 | 0.0008 |
| Anemia | 4.9 | 2.4,8.2 | 64.1 | 49.2,73.1 | 0.6 | 0.0155 |
| Jaundice | 4.7 | 2.4,7.7 | 59.9 | 42.4,70.3 | 0.4 | 0.0394 |

\*Egger's Bias plot statistical significance for asymmetry

**Fig 1.**
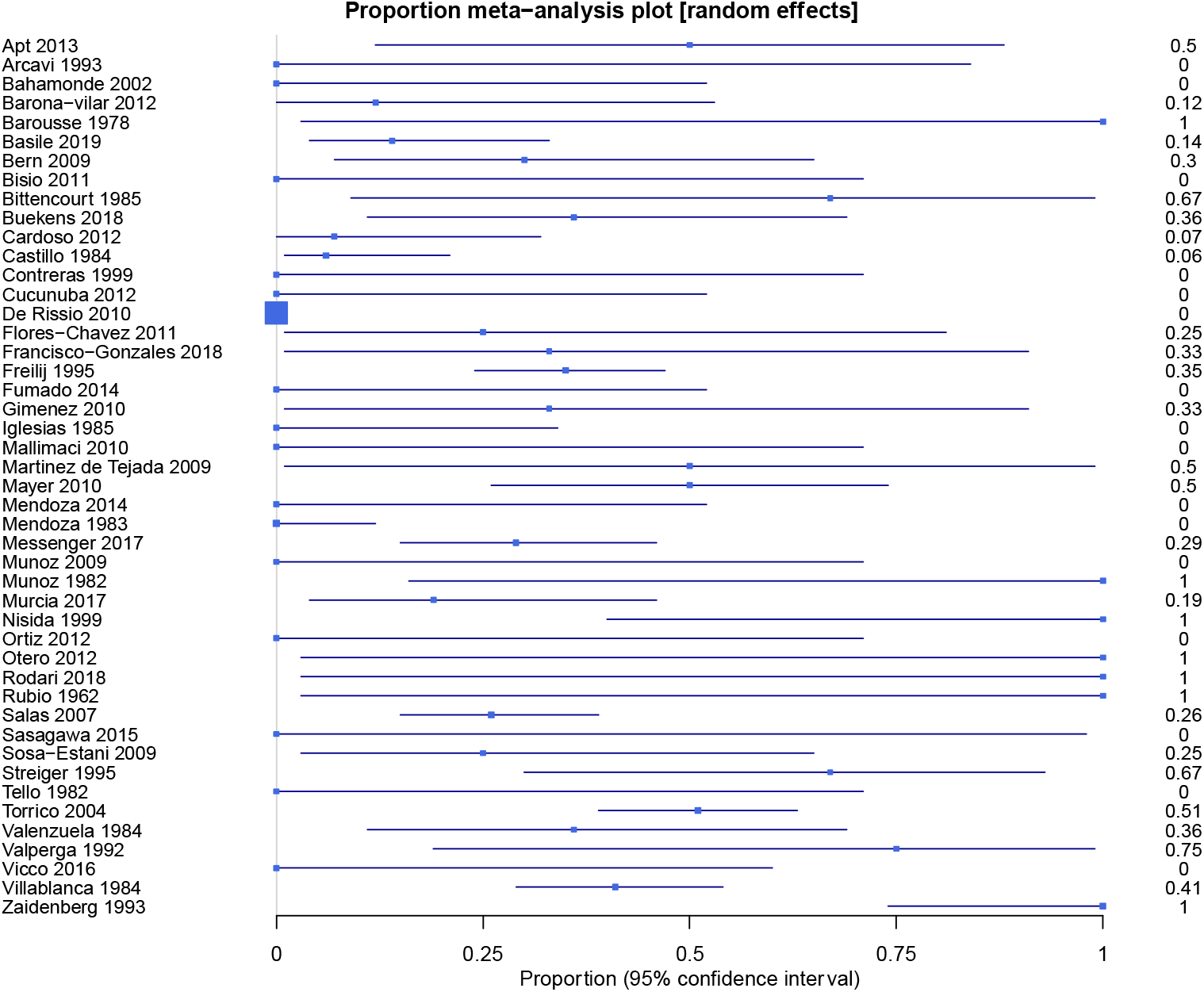
Morbidity Forest plot.

**Fig 2.**
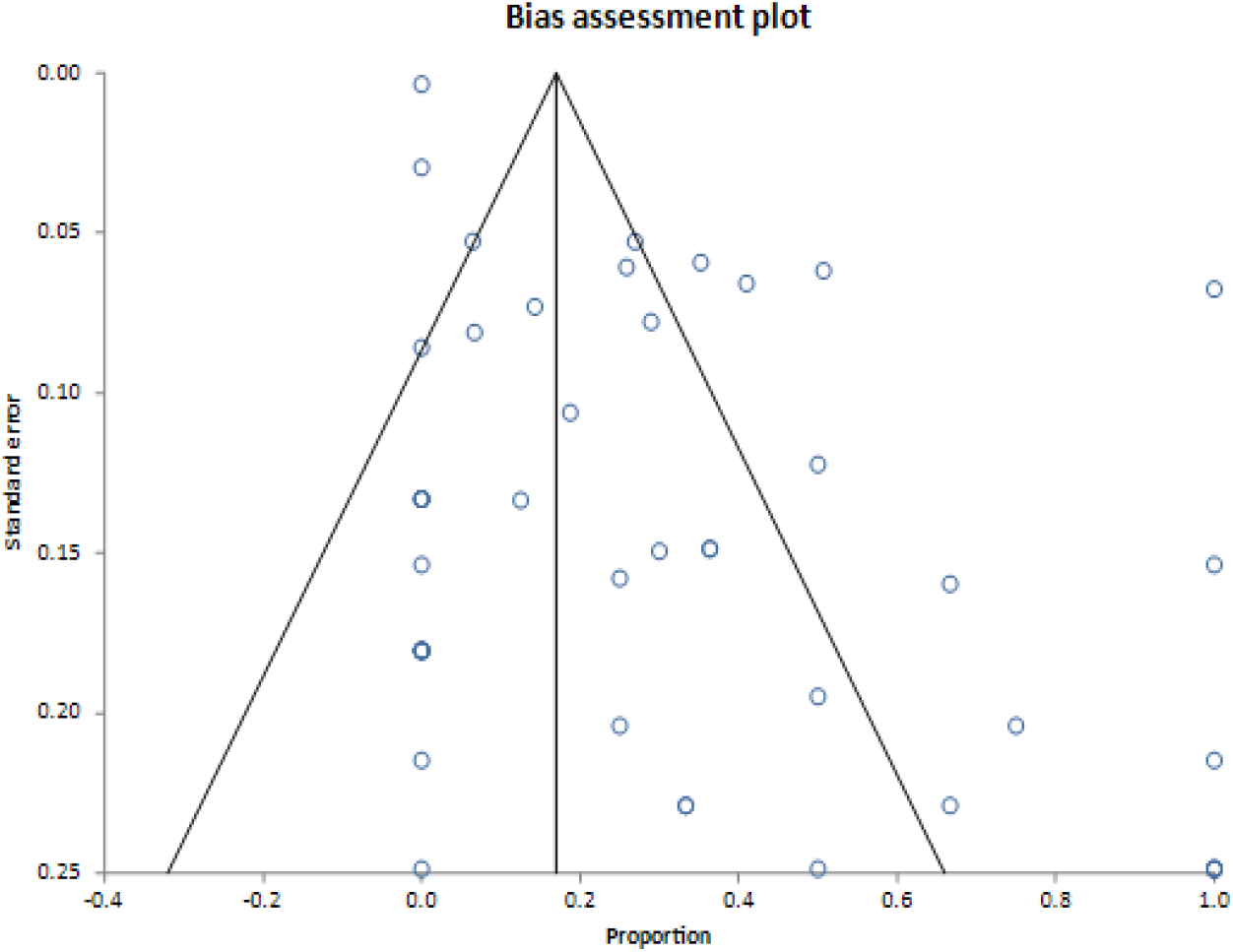
Morbidity Egger’s Bias plot.

The pooled proportion of cCD infected infants that died to all infected infants was 2.2% (95% CI = 1.3%, 3.5%) (**Fig 3**). The I^2^ inconsistency statistic was 9.6% (95% CI = 0% to 37.5%), suggesting between-study heterogeneity did not influence mortality. The 0.26 Egger’s bias statistic (**Fig 4**) was statistically significant (P = 0.0084), suggesting publication bias influenced results.

**Fig 3.**
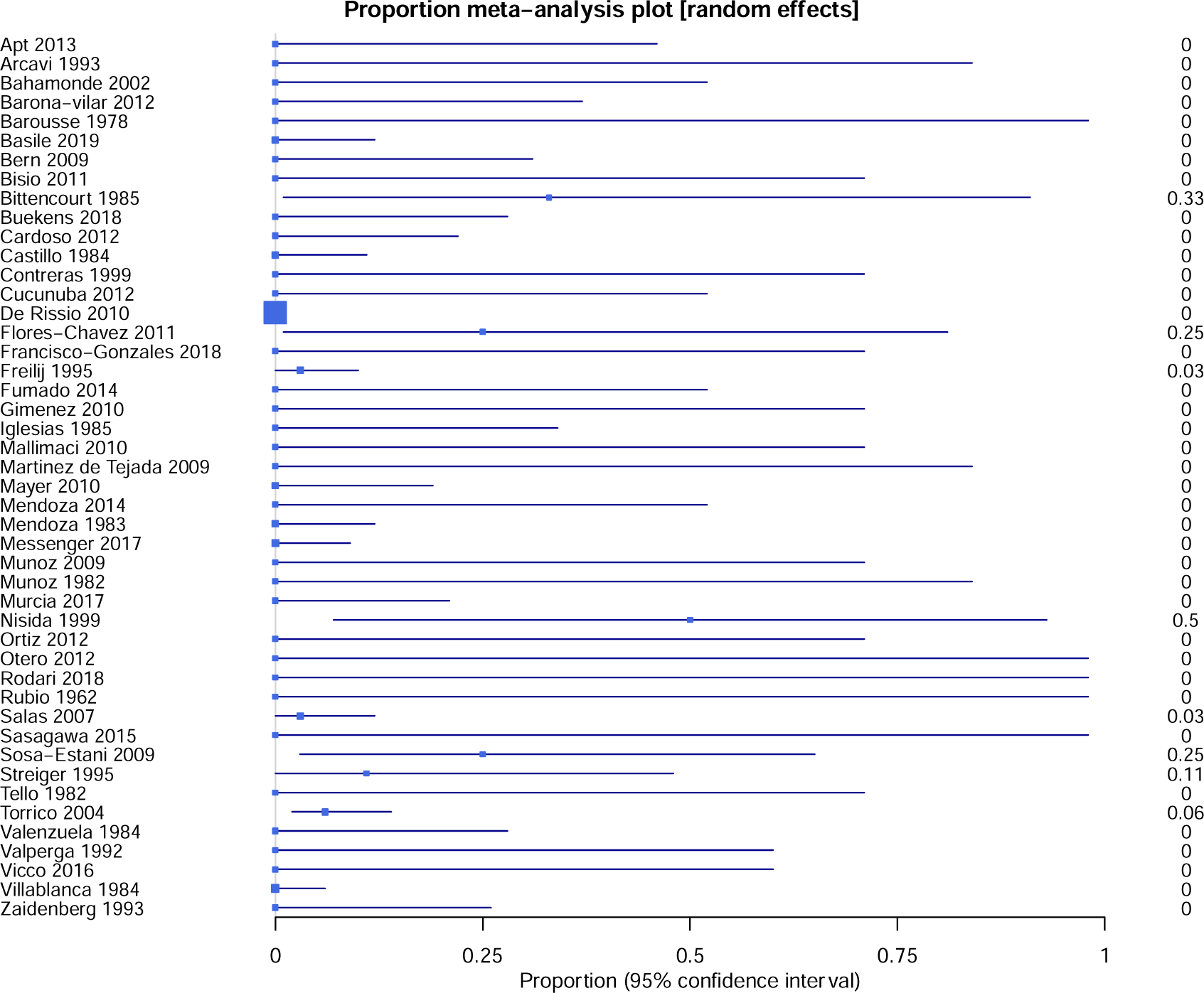
Morbidity Forest plot.

**Fig 4.**
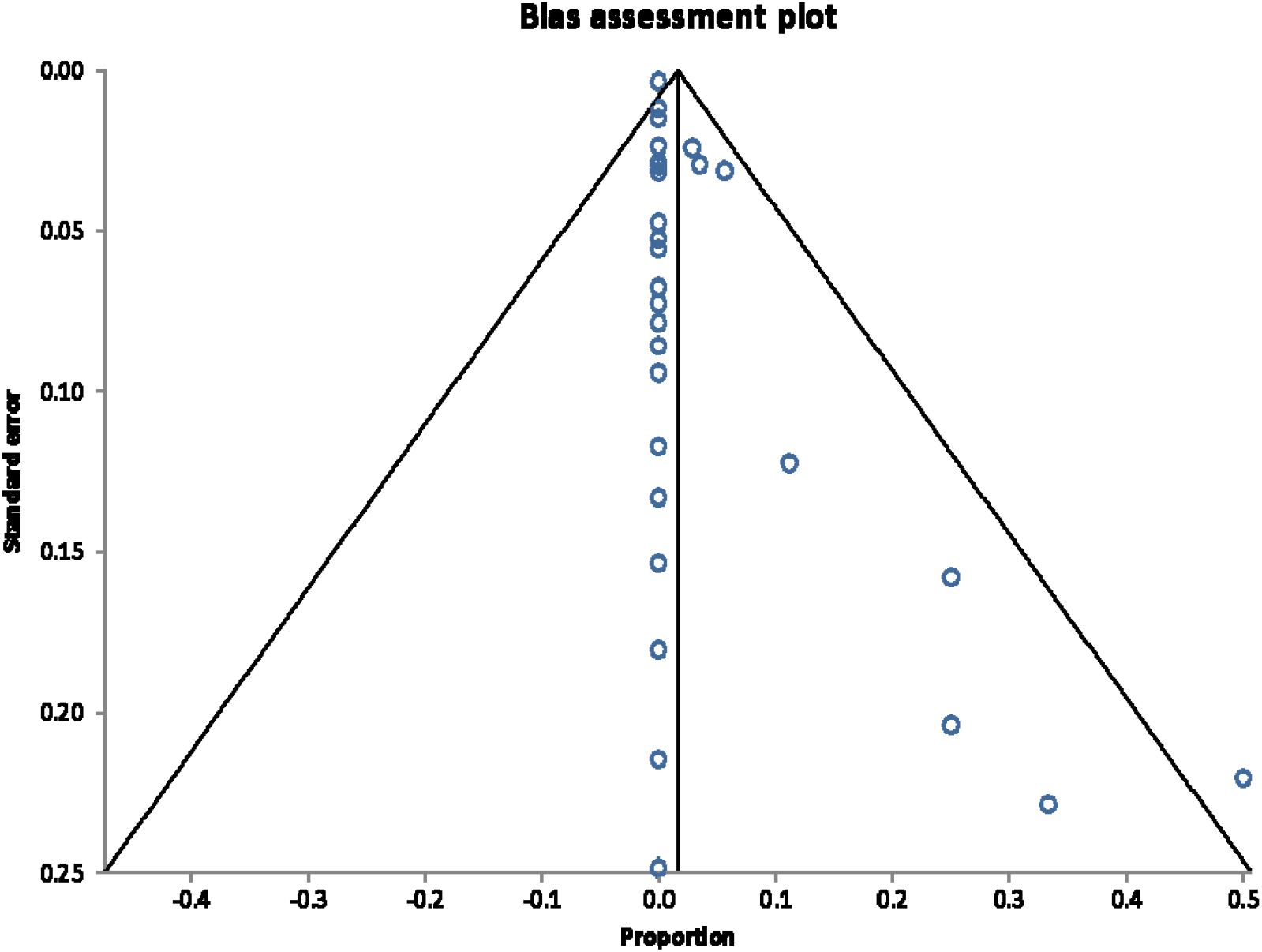
Morbidity Egger’s Bias plot.

### Subgroup analyses

Subgroup analysis results are in **Table 2**. Subgroup 1 analysed 45 studies with available information by whether gold standard diagnosis was used for cCD. The pooled proportion of symptomatic infants with cCD diagnosed with the gold standard was 18.7% (95% CI 6.1%, 36.1%), versus 32.5% (95% CI 23.0%, 42.9%) among infants diagnosed with an alternative.

Subgroup 2 analysed the proportion of symptomatic infants with cCD by geographic region in 47 studies. European studies (n=12) had a pooled proportion of 20.0% (95% CI 11.4%, 30.2%) versus 29.4% (95% CI 18.3%, 41.8%) in Latin American and Caribbean studies (n=35).

Subgroup 3 analysed the proportion of symptomatic infants with cCD by symptom to determine frequency of each symptom. Hepatosplenomegaly, reported as either hepatomegaly, splenomegaly, or hepatosplenomegaly, occurred most frequently, with a pooled proportion of 12.5% (95% CI 6.6%,19.9%). The following symptoms occurred the most frequently after hepatosplenomegaly: preterm birth with a pooled proportion of 6.0% (95% CI 3.3%, 9.5%), LBW 5.8% (95% CI 3.2%, 9.1%), anemia 4.9% (95% CI 2.4%, 8.2), and jaundice 4.7% (95% CI 2.4%, 7.7%).

## Discussion

### Main findings

Our primary meta-analysis of the proportion of symptomatic cCD infected infants to all cCD infected infants revealed a pooled proportion of 28.3% across 47 included studies. The pooled proportion of mortality cases among all cCD infected infants was estimated at 2.2%. Sensitivity analyses were conducted to determine robustness of results based on review decisions. Sources of heterogeneity were investigated based on infant characteristics and study characteristics across three subgroups. Detailed results and interpretations are described in **S6 File**.

### Interpretation

Our study expands on the body of work surrounding cCD and to our knowledge is the first to estimate its burden using an exhaustive search strategy that identified 47 studies for meta-analysis. Prior global estimates of the burden of cCD were likely underestimated given the influence of cited issues in diagnosing cCD on population-based data sources [18, 19]. Other estimations have been based on the results of individual observational studies [79, 80]. Our meta-analysis of observational studies allows for a more robust estimation of the burden of cCD in comparison to population-based data sources to describe the global burden of Chagas disease. A previous systematic review estimated that the pooled cCD transmission rate was 4.7% (95% CI: 3.9-5.6%) [3]. Our study suggests that of these cCD cases, 28.3% might present with morbidity and 2.2% with mortality. Compared to other congenital infections, about 10.0-30.0% of infants with congenital toxoplasmosis present with clinical symptoms at birth [81] and estimates from a study in Brazil suggest that 11.1% of congenital infections will result in fetal death [82]. In addition, 10.0-15.0% of infants born with congenital cytomegalovirus are symptomatic at birth with a mortality rate of <5% [83].

This study has raised concerns about the quality of studies that are conducted on cCD and their ability to attribute symptoms to the disease. Only two eligible studies compared symptoms in infected to non-infected mother-infant dyads [6, 60]. Torrico et al. revealed statistically significant increase in premature rupture of membranes and statistically significant decrease in birth weight and gestational age in infected dyads compared to non-infected dyads [6]. Similarly, Messenger et al. showed that *T*.*cruzi* infected infants were 2.7 times as likely to be low birthweight compared to non-infected infants (OR = 2.7, 95% CI 1.1, 5.8) [60]. Despite low risk of bias in these two studies, most other included studies were found to be moderate or high risk of bias. Coupled with a lack of comparison group, these studies have limited capability of attributing infected infants’ signs and symptoms to *T. cruzi* infection. Higher quality observational studies of cCD are needed.

There are various barriers to improving quality of cCD research. First, given that Chagas is defined as a neglected tropical disease (NTD) by the World Health Organization (WHO) and primarily affects impoverished populations, few resources have been dedicated to addressing the disease [84]. Disease control efforts historically have focused on vector control [85] historically leaving health systems unprepared to address cCD [86]. This is reflected in poor quality of studies published prior to major regional efforts in Latin America and the Caribbean (2017) [87]. The World Heart Federation has identified gaps in efforts to reduce cCD including ill-prepared healthcare personnel and lack of pregnancy screening programs [86]. The control strategies to close these gaps are cost-effective and reduce direct and indirect costs due to disease complications and death [88–90]. Despite this, the body of cCD literature still lacks in quality and further investment is needed.

We identified moderate to high risk of bias in over half of the included studies in reporting of results (71%), exposure and outcome measurement (65%), statistical methods (61%), and declaration of conflict and ethical statements (56%). Studies performed poorly in cCD diagnosis and reporting of these results, which has been cited as an issue due to limited access to and performance of the gold standard diagnostic algorithm, and the subsequent estimated 50% loss to follow-up of at-risk infants [3, 18]. In regard to outcome measurement, some studies only report signs and symptoms displayed, making it possible some may have been missed if studies did not explicitly evaluate for them. Furthermore, certain symptoms were not reported frequently enough to be analyzed such as intensive care unit (ICU) admission rate and low Apgar score. Four cases across three studies reported a low Apgar score (below 7 at 1 minute), and seven cases across three studies were admitted to the ICU. Low reporting frequency may be due to limitations in studies method of reporting; however, these symptoms are an important proxy for clinical severity. Furthermore, the proportion of infants presenting with low birth weight was 6.0%, lower than the rate in Latin America and the Caribbean (8.7%), North America, Europe, Australia, and New Zealand (7.0%), and globally (14.6%) [91]. This number is lower than expected and may be due to issues in outcome measurement and low-quality reporting of results. Given that previous literature has identified signs and symptoms of cCD through individual studies [2, 6–8, 10, 11, 79], the exhaustive list of signs and symptoms identified in this study can improve clinical surveillance and guide outcome measurement in future observational research.

The burden of cCD may increase as untreated children grow older and become chronic cases that may develop cardiac and/or gastrointestinal symptoms [18]. cCD is almost 100% curable in infants less than 1 year old and treatments are tolerated well [18]. In addition, treating infected women and girls before they bear children can prevent vertical transmission of *T. cruzi* [92, 93]. As such, our estimated proportion of 28.3% of cCD cases that present with symptoms may be preventable through increased screening and treatment. Despite this, an analysis of the 2010 Global Burden of Disease project data revealed that the decrease in Chagas’ burden of disease in DALYs was lower than that of other NTDs from 1990 to 2010 [94]. Given this burden is preventable, more investment in disease control and our understanding of its burden is needed.

### Strengths and limitations

This study has several strengths. First, to our knowledge there exists no other study that provides a pooled proportion of symptomatic cCD infected infants. This study employed a comprehensive search strategy, employed on databases that include those primarily focused on Latin American research, without language restrictions. Additionally, estimates produced were precise, as shown by narrow confidence intervals. The subgroup analysis focused on geographic region allowed for informed analyses of how this factor influences the proportion of symptomatic cCD infected infants. The mortality proportions estimates had low heterogeneity, suggesting studies are similar enough to combine and confidently interpret their results. The subgroup analysis of method of diagnosis informs how using a gold standard diagnosis influences the proportion of symptomatic cCD infected infants. Lastly, subgroup analysis by symptoms displayed provides further insight on the symptoms that are indicative of cCD in infants.

This study also has several limitations. First, grey literature was not searched, and given the statistical significance of the Egger’s bias estimate, this study is vulnerable to the effects of publication bias and ultimately its generalizability and validity. Additionally, most included studies did not compare morbidity or mortality in infected and non-infected mother-infant dyads. Without the comparison to an non-infected control group, this limits ability to associate signs and symptoms to cCD. The subgroup analysis of geographic region did not allow for disaggregation of results further than Latin American and Caribbean region due to a small sample size of studies from Mexico and Central America to analyse separately. Certain mortalities such as abortion and stillbirth may be underreported as these cases were only included in this analysis if the fetus has been diagnosed with Chagas disease post-mortem. Apgar scores were only collected at 1 minute as the majority of studies did not report scores at 5 minutes. Additionally, the majority of I^2^ estimates for morbidity proportions displayed considerable heterogeneity between studies, suggesting inconsistencies between studies are not due to chance alone and thus caution should be used when interpreting results. The risk of bias assessment revealed that overall, 34(72.3%) of included articles had a high risk of bias, 10 (21.3%) of articles had a moderate risk of bias, and only 3(6.4%) of articles had low risk of bias. This, in combination with a significant difference between the sensitivity analysis results excluding those studies with high risk of bias, suggests that the risk of bias influencing the results is high. Lastly, there was a large portion of studies with missing data for certain symptoms and missing values were assumed to be 0. Although this method likely meets the assumption that studies only reported symptoms that were displayed and all other values were zero, there is a chance this assumption was not met, and bias may have been introduced into these subgroup results due to this imputation.

## Conclusion

Among 47 included studies, the pooled proportion of symptomatic infections of cCD among all infected fetuses and infants was 28.3%; the pooled proportion of mortality for cCD among all cCD infected fetuses and infants was 2.2%. Caution should be used when interpreting estimated morbidity proportions, as there was considerable heterogeneity between studies.

Furthermore, sensitivity analyses revealed that excluding studies with a high risk of bias was significantly lower than the overall proportion (16.6%). Mortality proportions had low heterogeneity between studies and may be interpreted confidently. Studies comparing infected and non-infected mother-infant dyads are needed to determine the morbidity and mortality associated with cCD.

## Data Availability

All relevant data are within the manuscript and its Supporting Information files.

## Acknowledgements

The authors would like to thank Agustín Ciapponi and Luz Gibbons for their support and advice during this study.

## Contribution to authorship

**Conceptualization**: AFT, SM, PB, KPP, MEB, MC

**Study conduction, data collection and data analysis**: AFT, SM, PB, KPP, MEB, MC, DC

**Drafting of first manuscript**: AFT, SM

**Review and approval of final manuscript**: AFT, SM, PB, KP, MEB, MC, DC

## Supporting information

**S1 File**. Morbidity signs and symptoms of congenital Chagas disease

**S2 File**. Search strategy

**S3 File**. Hierarchy for consideration of full-text articles

**S4 File**. Summary of extracted data

**S5 File**. Risk of bias algorithms, summary within-domain risk of bias, and results

**S6 File**. Sensitivity analyses results and assessment of heterogeneity

**S1 Dataset**. Data extraction form

**S1 Table**. Congenital cases morbidity characteristics

**S2 Table**. Congenital cases mortality characteristics

