## Supplemental File 1 for "Estimation of the morbidity and mortality of congenital Chagas disease: a systematic review and meta-analysis"

**S1 File**. Morbidity signs and symptoms of congenital Chagas disease

- hepatomegaly
- splenomegaly
- respiratory distress syndrome
- neurologic signs (not including convulsions)
- anasarca
- petechiae
- low Apgar score (<7 at 1 minute)
- abnormal electrocardiographic findings
- anemia
- meningoencephalitis
- myocarditis
- congestive heart failure
- lesions in the digestive and/or central nervous system
- parasites in various tissues
- subependymal hemorrhage
- cardiomegaly
- premature rupture of the membrane (PROM)
- preterm birth
- low birth weight
- intra-uterine growth restriction
- small for gestational age
- neonatal ICU admission
