## Supplemental File 2 for "Estimation of the morbidity and mortality of congenital Chagas disease: a systematic review and meta-analysis"

**S2 File.** Search Strategy

LILACS (BVS-EN) 5-09-2019

Principio del formulario

| 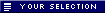 | 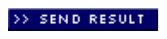 | 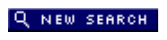 | 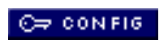 | 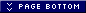<http://bases.bireme.br/cgi-bin/wxislind.exe/iah/online/#bottom> |
| --- | --- | --- | --- | --- |

| Database : | **LILACS** |
| --- | --- |
| Search on : | **(MH Chagas Disease OR Chagas OR Tiypanosom$ OR Trypanosom$ OR Cruzi OR T.Cruzi) AND (MH Infectious Disease Transmission, Vertical OR MH Pregnancy Complications, Infectious OR Vertical OR Fetomaternal OR Congenit$ OR Intrauterin$ OR Intra-Uterin$ OR MH Pregnancy OR Pregnan$ OR Embarazada$ OR Gravid$ OR MH Infant, Newborn OR Neonat$ OR Newborn$ OR Baby OR Babies OR Bebe OR Bebés OR Infant$ OR MH Fetus OR Fetus OR Feto OR Fetal OR MH Mothers OR Madre$ OR Mother$ OR Maes) [Words]** |
| References found : | **1007** [[refine](http://bases.bireme.br/cgi-bin/wxislind.exe/iah/online/#refine)] |

Final del formulario

EMBase (Elsevier) 5-09-2019

| No. | Query | Results |
| --- | --- | --- |
| #31 | #7 AND #30 | **2744** |
| #30 | #8 OR #9 OR #10 OR #11 OR #12 OR #13 OR #14 OR #15 OR #16 OR #17 OR #18 OR #19 OR #20 OR #21 OR #22 OR #23 OR #24 OR #25 OR #26 OR #27 OR #28 OR #29 | **2473573** |
| #29 | maternal:ti,ab | **306723** |
| #28 | mother*:ti,ab | **266877** |
| #27 | 'mother'/exp | **135154** |
| #26 | fetus:ti,ab | **82979** |
| #25 | fetal:ti,ab | **298553** |
| #24 | 'fetus'/exp | **198980** |
| #23 | infant*:ti,ab | **475549** |
| #22 | babies:ti,ab | **50145** |
| #21 | baby:ti,ab | **53168** |
| #20 | 'newly born':ti,ab | **1427** |
| #19 | 'new born':ti,ab | **5180** |
| #18 | newborn*:ti,ab | **198437** |
| #17 | neonat*:ti,ab | **331872** |
| #16 | 'newborn'/exp | **579229** |
| #15 | pregnan*:ti,ab | **636682** |
| #14 | 'pregnancy'/exp | **745344** |
| #13 | intrauterine:ti,ab | **68037** |
| #12 | congenital:ti,ab | **300313** |
| #11 | fetomaternal:ti,ab | **2279** |
| #10 | 'mother to child':ti,ab | **7540** |
| #9 | vertical:ti,ab | **106213** |
| #8 | 'vertical transmission'/exp | **14115** |
| #7 | #1 OR #2 OR #3 OR #4 OR #5 OR #6 | **48834** |
| #6 | t.cruzi:ti,ab | **9291** |
| #5 | cruzi:ti,ab | **16453** |
| #4 | trypanosom*:ti,ab | **35668** |
| #3 | 'trypanosomiasis'/exp | **26385** |
| #2 | chagas*:ti,ab | **16435** |
| #1 | 'chagas disease'/exp | **1609** |

PubMed 5-09-2019

| Search | Query | Items found | Time |
| --- | --- | --- | --- |
| [#32](https://www.ncbi.nlm.nih.gov/pubmed/advanced) | Search **(#7 AND #31)** | [2341](https://www.ncbi.nlm.nih.gov/pubmed/?cmd=HistorySearch&querykey=32) | 14:13:17 |
| [#31](https://www.ncbi.nlm.nih.gov/pubmed/advanced) | Search **(#8 OR #9 OR #10 OR #11 OR #12 OR #13 OR #14 OR #15 OR #16 OR #17 OR #18 OR #19 OR #20 OR #21 OR #22 OR #23 OR #24 OR #25 OR #26 OR #27 OR #28 OR #29 OR #30)** | [2200274](https://www.ncbi.nlm.nih.gov/pubmed/?cmd=HistorySearch&querykey=31) | 14:09:12 |
| [#30](https://www.ncbi.nlm.nih.gov/pubmed/advanced) | Search **Maternal[tiab]** | [244275](https://www.ncbi.nlm.nih.gov/pubmed/?cmd=HistorySearch&querykey=30) | 14:08:54 |
| [#29](https://www.ncbi.nlm.nih.gov/pubmed/advanced) | Search **Mother*[tiab]** | [209149](https://www.ncbi.nlm.nih.gov/pubmed/?cmd=HistorySearch&querykey=29) | 14:08:42 |
| [#28](https://www.ncbi.nlm.nih.gov/pubmed/advanced) | Search **Mothers[Mesh]** | [40730](https://www.ncbi.nlm.nih.gov/pubmed/?cmd=HistorySearch&querykey=28) | 14:08:21 |
| [#27](https://www.ncbi.nlm.nih.gov/pubmed/advanced) | Search **Fetus[tiab]** | [67722](https://www.ncbi.nlm.nih.gov/pubmed/?cmd=HistorySearch&querykey=27) | 14:08:13 |
| [#26](https://www.ncbi.nlm.nih.gov/pubmed/advanced) | Search **Fetal[tiab]** | [236692](https://www.ncbi.nlm.nih.gov/pubmed/?cmd=HistorySearch&querykey=26) | 14:08:04 |
| [#25](https://www.ncbi.nlm.nih.gov/pubmed/advanced) | Search **Fetus[Mesh]** | [154644](https://www.ncbi.nlm.nih.gov/pubmed/?cmd=HistorySearch&querykey=25) | 14:07:56 |
| [#24](https://www.ncbi.nlm.nih.gov/pubmed/advanced) | Search **Infant*[tiab]** | [431221](https://www.ncbi.nlm.nih.gov/pubmed/?cmd=HistorySearch&querykey=24) | 14:07:48 |
| [#23](https://www.ncbi.nlm.nih.gov/pubmed/advanced) | Search **Babies[tiab]** | [35529](https://www.ncbi.nlm.nih.gov/pubmed/?cmd=HistorySearch&querykey=23) | 14:07:36 |
| [#22](https://www.ncbi.nlm.nih.gov/pubmed/advanced) | Search **Baby[tiab]** | [36746](https://www.ncbi.nlm.nih.gov/pubmed/?cmd=HistorySearch&querykey=22) | 14:07:26 |
| [#21](https://www.ncbi.nlm.nih.gov/pubmed/advanced) | Search **Newly Born*[tiab]** | [1101](https://www.ncbi.nlm.nih.gov/pubmed/?cmd=HistorySearch&querykey=21) | 14:07:18 |
| [#20](https://www.ncbi.nlm.nih.gov/pubmed/advanced) | Search **New Born*[tiab]** | [4329](https://www.ncbi.nlm.nih.gov/pubmed/?cmd=HistorySearch&querykey=20) | 14:07:10 |
| [#19](https://www.ncbi.nlm.nih.gov/pubmed/advanced) | Search **Newborn*[tiab]** | [170189](https://www.ncbi.nlm.nih.gov/pubmed/?cmd=HistorySearch&querykey=19) | 14:07:02 |
| [#18](https://www.ncbi.nlm.nih.gov/pubmed/advanced) | Search **Neonat*[tiab]** | [256533](https://www.ncbi.nlm.nih.gov/pubmed/?cmd=HistorySearch&querykey=18) | 14:06:55 |
| [#17](https://www.ncbi.nlm.nih.gov/pubmed/advanced) | Search **Infant, Newborn[Mesh]** | [589320](https://www.ncbi.nlm.nih.gov/pubmed/?cmd=HistorySearch&querykey=17) | 14:06:47 |
| [#16](https://www.ncbi.nlm.nih.gov/pubmed/advanced) | Search **Pregnan*[tiab]** | [501050](https://www.ncbi.nlm.nih.gov/pubmed/?cmd=HistorySearch&querykey=16) | 14:06:38 |
| [#15](https://www.ncbi.nlm.nih.gov/pubmed/advanced) | Search **Pregnancy[Mesh]** | [868799](https://www.ncbi.nlm.nih.gov/pubmed/?cmd=HistorySearch&querykey=15) | 14:06:31 |
| [#14](https://www.ncbi.nlm.nih.gov/pubmed/advanced) | Search **Intrauterine[tiab]** | [50447](https://www.ncbi.nlm.nih.gov/pubmed/?cmd=HistorySearch&querykey=14) | 14:06:23 |
| [#13](https://www.ncbi.nlm.nih.gov/pubmed/advanced) | Search **Congenital[tiab]** | [238682](https://www.ncbi.nlm.nih.gov/pubmed/?cmd=HistorySearch&querykey=13) | 14:06:13 |
| [#12](https://www.ncbi.nlm.nih.gov/pubmed/advanced) | Search **Fetomaternal[tiab]** | [1686](https://www.ncbi.nlm.nih.gov/pubmed/?cmd=HistorySearch&querykey=12) | 14:06:02 |
| [#11](https://www.ncbi.nlm.nih.gov/pubmed/advanced) | Search **"Mother to Child"[tiab]** | [6212](https://www.ncbi.nlm.nih.gov/pubmed/?cmd=HistorySearch&querykey=11) | 14:05:54 |
| [#10](https://www.ncbi.nlm.nih.gov/pubmed/advanced) | Search **Vertical[tiab]** | [94683](https://www.ncbi.nlm.nih.gov/pubmed/?cmd=HistorySearch&querykey=10) | 14:05:44 |
| [#9](https://www.ncbi.nlm.nih.gov/pubmed/advanced) | Search **Pregnancy Complications, Infectious[Mesh]** | [43498](https://www.ncbi.nlm.nih.gov/pubmed/?cmd=HistorySearch&querykey=9) | 14:05:35 |
| [#8](https://www.ncbi.nlm.nih.gov/pubmed/advanced) | Search **Infectious Disease Transmission, Vertical[Mesh]** | [15229](https://www.ncbi.nlm.nih.gov/pubmed/?cmd=HistorySearch&querykey=8) | 14:05:24 |
| [#7](https://www.ncbi.nlm.nih.gov/pubmed/advanced) | Search **(#1 OR #2 OR #3 OR #4 OR #5 OR #6)** | [42743](https://www.ncbi.nlm.nih.gov/pubmed/?cmd=HistorySearch&querykey=7) | 13:52:22 |
| [#6](https://www.ncbi.nlm.nih.gov/pubmed/advanced) | Search **T.Cruzi[tiab]** | [8241](https://www.ncbi.nlm.nih.gov/pubmed/?cmd=HistorySearch&querykey=6) | 13:52:07 |
| [#5](https://www.ncbi.nlm.nih.gov/pubmed/advanced) | Search **Cruzi[tiab]** | [14735](https://www.ncbi.nlm.nih.gov/pubmed/?cmd=HistorySearch&querykey=5) | 13:52:00 |
| [#4](https://www.ncbi.nlm.nih.gov/pubmed/advanced) | Search **Trypanosom*[tiab]** | [34203](https://www.ncbi.nlm.nih.gov/pubmed/?cmd=HistorySearch&querykey=4) | 13:51:51 |
| [#3](https://www.ncbi.nlm.nih.gov/pubmed/advanced) | Search **Trypanosomiasis[Mesh]** | [21668](https://www.ncbi.nlm.nih.gov/pubmed/?cmd=HistorySearch&querykey=3) | 13:51:40 |
| [#2](https://www.ncbi.nlm.nih.gov/pubmed/advanced) | Search **Chagas*[tiab]** | [14149](https://www.ncbi.nlm.nih.gov/pubmed/?cmd=HistorySearch&querykey=2) | 13:51:28 |
| [#1](https://www.ncbi.nlm.nih.gov/pubmed/advanced) | Search **Chagas Disease[Mesh]** | [12756](https://www.ncbi.nlm.nih.gov/pubmed/?cmd=HistorySearch&querykey=1) | 13:51:16 |

CINAHL (EBSCO) 10-09-2019

| **#** | **Query** | **Results** |
| --- | --- | --- |
| S31 | S6 AND S30 | 111 |
| S30 | S7 OR S8 OR S9 OR S10 OR S11 OR S12 OR S13 OR S14 OR S15 OR S16 OR S17 OR S18 OR S19 OR S20 OR S21 OR S22 OR S23 OR S24 OR S25 OR S26 OR S27 OR S28 OR S29 | 284,146 |
| S29 | TI Maternal OR AB Maternal | 54,612 |
| S28 | TI Mother* OR AB Mother* | 59,971 |
| S27 | (MH "Mothers+") | 24,147 |
| S26 | TI Fetal OR AB Fetal | 28,478 |
| S25 | TI Fetus OR AB Fetus | 13,106 |
| S24 | TI Infant* OR AB Infant* | 71,104 |
| S23 | (MH "Fetus+") | 12,688 |
| S22 | TI Babies OR AB Babies | 19,250 |
| S21 | TI Baby OR AB Baby | 19,590 |
| S20 | TI "Newly Born" OR AB "Newly Born" | 98 |
| S19 | TI "New Born" OR AB "New Born" | 258 |
| S18 | TI Intrauterine OR AB Intrauterine | 7,365 |
| S17 | TI Newborn* OR AB Newborn* | 20,494 |
| S16 | TI Neonat* OR AB Neonat* | 43,525 |
| S15 | (MH "Infant, Newborn+") | 71,346 |
| S14 | TI Pregnan* OR AB Pregnan* | 86,676 |
| S13 | (MH "Pregnancy+") | 102,158 |
| S12 | TI Congenital OR AB Congenital | 24,767 |
| S11 | TI Fetomaternal OR AB Fetomaternal | 177 |
| S10 | TI "Mother to Child" OR AB "Mother to Child" | 3,123 |
| S9 | TI Vertical OR AB Vertical | 12,257 |
| S8 | (MH "Pregnancy Complications, Infectious+") | 3,223 |
| S7 | (MM "Disease Transmission, Vertical") | 1,882 |
| S6 | S1 OR S2 OR S3 OR S4 OR S5 | 953 |
| S5 | TI T.Cruzi OR AB T.Cruzi | 2 |
| S4 | TI Cruzi OR AB Cruzi | 363 |
| S3 | TI Trypanosom* OR AB Trypanosom* | 587 |
| S2 | TI Chagas* OR AB Chagas* | 574 |
| S1 | (MM "Trypanosomiasis") | 432 |

Academic Search Complete (EBSCO) 10-09-2019

| # | Query | Results |
| --- | --- | --- |
| S29 | S7 AND S28 | 815 |
| S28 | S8 OR S9 OR S10 OR S11 OR S12 OR S13 OR S14 OR S15 OR S16 OR S17 OR S18 OR S19 OR S20 OR S21 OR S22 OR S23 OR S24 OR S25 OR S26 OR S27 | 805,707 |
| S27 | TI Fetal OR AB Fetal | 85,717 |
| S26 | TI Maternal AND AB Maternal | 30,583 |
| S25 | DE "MOTHERS" | 16,248 |
| S24 | TI Mother* AND AB Mother* | 34,134 |
| S23 | TI Fetus OR AB Fetus | 38,252 |
| S22 | DE "FETUS" | 7,427 |
| S21 | TI Infant* OR AB Infant* | 170,794 |
| S20 | TI Babies OR AB Babies | 77,497 |
| S19 | TI Baby OR AB Baby | 77,497 |
| S18 | TI "Newly Born" OR AB "Newly Born" | 792 |
| S17 | TI "New Born" OR AB "New Born" | 1,441 |
| S16 | TI Newborn* OR AB Newborn* | 52,947 |
| S15 | TI Neonat* OR AB Neonat* | 99,560 |
| S14 | TI Pregnan* OR AB Pregnan* | 219,231 |
| S13 | DE "PREGNANCY" | 72,275 |
| S12 | TI Intrauterine OR AB Intrauterine | 19,564 |
| S11 | TI Congenital OR AB Congenital | 69,798 |
| S10 | TI Fetomaternal OR AB Fetomaternal | 635 |
| S9 | TI "Mother to Child" OR AB "Mother to Child" | 6,957 |
| S8 | TI Vertical OR AB Vertical | 154,683 |
| S7 | S1 OR S2 OR S3 OR S4 OR S5 OR S6 | 14,873 |
| S6 | TI T.Cruzi OR AB T.Cruzi | 55 |
| S5 | TI Cruzi OR AB Cruzi | 5,525 |
| S4 | TI Trypanosom* OR AB Trypanosom* | 12,150 |
| S3 | DE "TRYPANOSOMIASIS" | 1,395 |
| S2 | TI Chagas* OR AB Chagas* | 5,540 |
| S1 | DE "CHAGAS' disease" | 2,695 |
