## Supplemental File 3 for "Estimation of the morbidity and mortality of congenital Chagas disease: a systematic review and meta-analysis"

**S3 File.** Hierarchy for consideration of full-text articles

**Reviewers’ reason for exclusion rationale:**

The following list is in order of what reviewers used to evaluate an article’s eligibility for data extraction during the full text review. This ordered list was made to identify the appropriate reason for exclusion for articles. See list below:

1. Ineligible study design: No original data
2. Ineligible study design: Other study designs outlined as ineligible
3. Ineligible study population
4. No congenital transmission documented
5. No morbidity/mortality described
6. Denominator cannot be determined

Other reasons for exclusion that are not documented in this list were also used by reviewers. These reasons for exclusion are listed below:

- Duplicates (excluded)
- Duplicates (included)
- No full text

The description of each reason for exclusion can be found in the following section:

**Descriptions of reasons for exclusion:**

1. **No morbidity/mortality described**: Article did not provide any information on the congenital cases’ clinical symptoms, thus reviewers had no way of knowing if congenital morbidity/mortality was seen among patients.
2. **Duplicates (excluded)**: Articles that are a duplicate of an excluded article in any other category.
3. **Duplicates (included)**: Articles that are a duplicate of an included article
4. **Ineligible study population**: The study population did not meet the inclusion criteria (e.g. a mixed cohort of children and neonates where crude data for neonates was not provided, a cohort of only children and not neonates, etc.)
5. **Ineligible study design**: The study design did not meet the inclusion criteria. Inclusion criteria specify that this review will exclude case reports, case series, any studies that do not include original data, or case-control studies that incidence estimates cannot be calculated for the neonatal population.
6. **No full text**: The full text of this article could not be retrieved after contacting Tulane SPHTM’s medical librarian and IECS’s medical librarian.
7. **No congenital transmission documented**: The article did not document any cases of congenital transmission among neonates.
8. **Denominator could not be determined**: The denominator for the total number of congenitally infected infants could not be determined.
