## Supplemental File 4 for "Estimation of the morbidity and mortality of congenital Chagas disease: a systematic review and meta-analysis"

**S4 File.** Summary of extracted data

Extracted study characteristics were the article ID in Covidence, authors, year of publication, region, country, entity, city, country endemicity, location endemicity, study setting, study design, year of dataset, study objective, and source population. Extracted maternal characteristics were the number of participants and the number of those that completed follow-up, mothers’ nationality, residency type (urban, rural, mixed), socioeconomic status, the number of Chagas cases, and any additional reported characteristics. Extracted infant characteristics were the number of participants and the number of those that completed follow-up, any study participant characteristics, and birth complications not related to Chagas. Diagnostic information for Chagas disease in mothers and cCD in infants included diagnostic test(s) used, diagnostic sample(s) used, and timing of diagnostic test(s). In addition to morbidity symptoms, the total number of congenital cases, the number of asymptomatic and symptomatic cases, and any other reported symptoms not previously listed were further extracted. Mortality at birth to six days, seven to 27 days, and 28 days to one year were also extracted from each study.
