## Supplemental File 5 for "Estimation of the morbidity and mortality of congenital Chagas disease: a systematic review and meta-analysis"

**S5 File.** Risk of bias algorithms, summary within-domain risk of bias, and results

**Algorithm**

EXPOSURE = congenital Chagas diagnosed by gold standard method

OUTCOME = symptoms of Chagas infection

*Exposure assessed through gold standard method*

The gold standard for diagnosis of congenital Chagas disease is **parasitological examination using the microhematocrit or microstrout** testing methods (Carlier et al., 2019). This test **should be performed at birth, and if negative the test should be repeated again one month later** (Carlier et al., 2019). **Serological tests should be performed if there was no previous screening of the neonate or the parasitological and/or molecular test results were negative** (Carlier et al., 2019). **Serological assays can be performed at 10 months of age, when maternal antibodies have waned in the infant**, (Carlier et al., 2019)

*Was loss to follow-up after baseline 20% or less?*

- 20% or less **LOW** risk of bias
- 21% - 30% **PARTIAL** risk of bias
- 31% or more **HIGH** risk of bias

*Descriptive data: characteristics of study participants (e.g. demographic, clinical, social) and information on exposure, indicate the number of participants with missing data for each variable of
interest, summarize follow-up time (e.g. average and total amount)*

- Characteristics = those that are outlined in our data extraction form.
  - Data extraction form -
    - Maternal characteristics:
      - Participants Nationality
      - Current Residence (Urban/Rural/Mixed)
      - Participants SES
      - Other Reported Characteristics
      - # Chagas cases
      - Diagnostic Test Used
      - Diagnostic Sample Used
      - Timing of Diagnostic
    - Congenital case characteristics:
      - Study Participant Characteristics
      - Birth Complications (Not Chagas Related)
  - If study includes mothers and infants
    - 7-10 **LOW** risk of bias
    - 4-6 **PARTIAL** risk of bias
    - 0-3 **HIGH** risk of bias
  - If study includes only infants
    - 2 **LOW** risk of bias
    - 1 **PARTIAL** risk of bias
    - 0 **HIGH** risk of bias

Scenarios where N/A was used:

- Methods for measuring exposure and outcome variables:
  - Was loss to follow-up after baseline 20% or less?
    - In retrospective case-control and cross-sectional studies, this was counted as N/A
- Methods to control confounding:
  - Only used in scenarios where measures of association were calculated
- Statistical methods:
  - Describe all statistical methods, including those used to control for confounding
    - If confounding was not measured, and studies were only reporting crude descriptive statistics, this was marked as N/A
  - *Cohort study—Explain how loss to follow-up was addressed
    Case-control study—Explain how matching of cases and controls was addressed
    Cross-sectional study—Describe analytical methods taking account of sampling strategy*
    - If there was no loss to follow-up
    - If there was no matching

**Summary within-domain risk of bias**

|  | **Methods for selecting study participants** | **Methods for measuring exposure and outcome variables** | **Methods to control confounding** | **Reporting of results** | **Statistical methods** | **Declaration of conflict and ethical statements** |
| --- | --- | --- | --- | --- | --- | --- |
| **Apt-2013** | / | + |  | / | + | + |
| **Arcavi-1993** | + | / |  | + | - | - |
| **Azogue-1991** | + | + |  | / |  | / |
| **Bahamonde-2002** | + | + |  | / | - | - |
| **Barona-Vilar-2012** | + | + |  | / | + | + |
| **Barousse-1978** | + | / |  | / | / | / |
| **Basile-2019** | + | / | + | + | / | + |
| **Bern-2009** | + | / |  | / | / | + |
| **Bisio-2011** | + | / |  | / | / | + |
| **Bittencourt-1985** | / | / |  | + | - | - |
| **Buekens-2018** | + | + |  | + | / | + |
| **Cardoso-2012** | + | / |  | / | / | + |
| **Castillo-1984** | / | - |  | - | - | / |
| **Contreras-1999** | / | / |  | - | - | - |
| **Cucunuba-2012** | / | - |  | / | - | - |
| **deRissio-2010** | + | / |  | + | + | + |
| **Florez-Chavez-2011** | / | / |  | / | + | / |
| **Francisco-Gonzalez-2019** | / | + |  | / | + | + |
| **Freiliji-1995** | / | / |  | / | + | - |
| **Fumado-2014** | + | / |  | / | - | - |
| **Gimenez-2010** | / | / |  | / | - | - |
| **Iglesias-1985** | / | - |  | / | - | - |
| **Mallimaci-2010** | / | + |  | + | + | + |
| **Martinez de Tejada-2009** | + | / |  | / | - | - |
| **Mayer-2010** | + | + |  | / | + | + |
| **Mendoza-2014** | / | + |  | / | - | - |
| **Mendoza-1983** | + | / |  | + |  | / |
| **Messenger-2017** | + | / | + | + | + | + |
| **Munoz-2009** | + | + | + | + | / | + |
| **Munoz-1982** | + | + |  | / | - | / |
| **Murcia-2017** | / | / |  | - | + | + |
| **Nisida-1999** | / | / |  | / | - | / |
| **Ortiz-2012** | - | / |  | / | - | + |
| **Otero-2012** | + | + |  | + |  | / |
| **Rodari-2018** | / | + |  | / | / | / |
| **Rubio-1962** | - | / |  | / | - | - |
| **Salas-2007** | + | - | + | / | / | + |
| **Sasagawa-2015** | / | + |  | + | / | + |
| **Sosa-Estani-2009** | + | / |  | / | + | + |
| **Streiger-1995** | + | / |  | / | - | / |
| **Tello-1982** | + | / |  | / | - | / |
| **Torrico-2004** | / | / |  | - | + | + |
| **Valenzuela-1984** | / | - |  | / |  | / |
| **Valperga-1992** | / | + |  | + |  | - |
| **Vicco-2016** | / | + |  | + | / | + |
| **Villablanca-1984** | / | - |  | / |  | / |
| **Zaidenberg-1993** | / | / |  | / | - | - |

**Results**

Within domain 1, 23(47.9%) articles had low risk of bias, 22(45.8%) had moderate, and 2(4.2%) had high risk. In Domain 2, 16(33.3%), 25(52.1%), and 6(12.5%) articles had low, moderate, and high risks of bias, respectively. Domain 3 had 4(8.3%) articles with low risk of bias; the remaining 43 articles were non-applicable within this domain. Domain 4 had 13(27.1%), 30(62.5%), and 4(8.3%) articles with low, moderate, and high risks of bias, respectively. Domain 5 had 12(25.0%) low, 11(22.9%) moderate, and 18(37.5%) high risks of bias; the remaining 6 articles were non-applicable. Domain 6 had 20(41.7%) articles with low risk of bias, 13(27.1%) moderate, and 14(29.2%) high. **Fig 5** summarizes the within-domain risk of bias assessment. Overall, 34(72.3%) articles were determined to have high risk of bias, 10(21.3%) were moderate, and 3(6.4%) were low.

**Fig 5. Within-domain Risk of Bias Summary.**

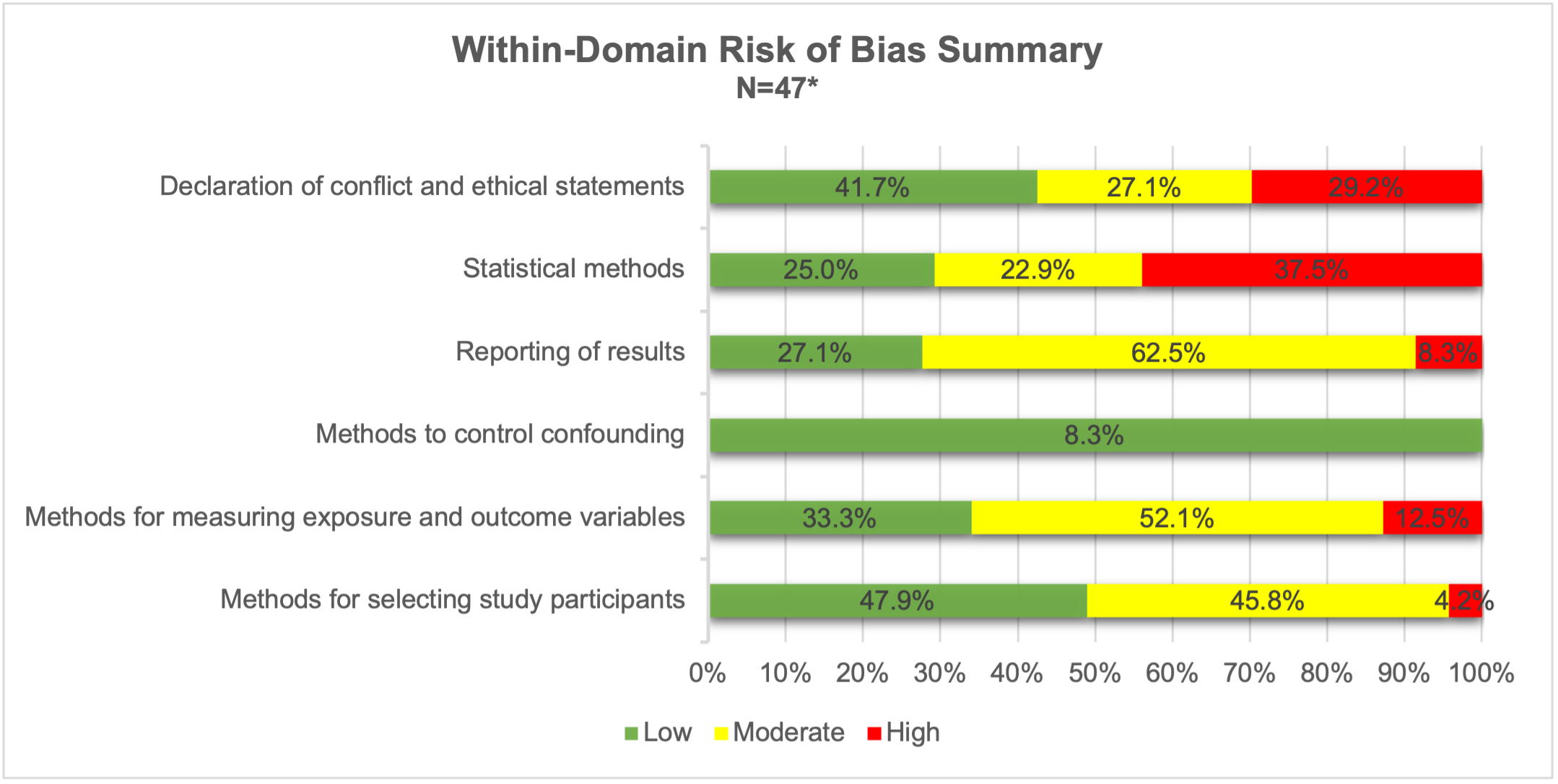

**^*^**Does not include percentage of N/A values within domains therefore totals for each category will not equate to 100%
