## Supplemental File 6 for "Estimation of the morbidity and mortality of congenital Chagas disease: a systematic review and meta-analysis"

**S6 File.** Sensitivity analyses results and assessment of heterogeneity

Results from sensitivity analyses are found in **Table 3**. Excluding studies with high risk of bias (n=13), the pooled proportion of infants symptomatic for cCD to all cCD infected infants was 16.9% (95% CI 5.3%, 33.5%). Excluding mothers not diagnosed by WHO guidelines (n=28), the pooled proportion was 31.2% (95% CI 17.5%, 47.0%). Using the Miller back-transformation, the pooled proportion of infants symptomatic for cCD to all cCD infected infants was 23.2% (95% CI 12.1%, 35.8%) and the pooled proportion of mortality was 0.0% (95% CI = 0.0%,0.0%).

**Table 3. Sensitivity Analyses**

|  | **Pooled Proportion %** | 95% CI % | **I^2^ (%)** | 95% CI % | **Egger's Bias** | P-Value^*^ |
| --- | --- | --- | --- | --- | --- | --- |
| ***Primary Analyses (N=47)*** |  |  |  |  |  |  |
| Morbidity | 28.3 | 19.0,38.5 | 88.6 | 86.0,90.5 | 2.5 | <0.0001 |
| Mortality | 2.2 | 1.3,3.5 | 9.6 | 0.0,37.5 | 0.3 | 0.0084 |
| ***Sensitivity Analyses (N=47)*** |  |  |  |  |  |  |
| Low/Moderate Risk of Bias (n=13) | 16.9 | 5.3,33.5 | 91.4 | 89.2,93.0 | 1.9 | 0.0078 |
| WHO Recommended Maternal Diagnosis (n=28) | 31.2 | 17.5,47.0 | 91.2 | 87.3,93.5 | 2.7 | 0.0003 |
| ***Ad-Hoc Miller Transformation (N=47)*** |  |  |  |  |  |  |
| Morbidity | 23.2 | 12.1,35.8 | 88.0 | 86.0,90.5 | 2.5 | <0.0001 |
| Mortality | 0.0 | 0.0,0.0 | 7.8 | 0.0,36.0 | 0.3 | 0.0085 |

*^*^Egger's Bias plot statistical significance for asymmetry*

Our meta-analysis was performed using the Freeman-Tukey double arcsine method and applying Stuart-Ord inverse variance weights to transformed proportions. Using the Miller back-transformation, the estimated summary frequency decreased to 23.2% for morbidity and 0.0% for mortality. For morbidity, the summary frequency differs enough to conclude results are not robust and may be influenced by the variance transformation used. For mortality, however, results are robust. Differences in these values may be attributable to the fact there is high heterogeneity between studies for morbidity but may not have been a factor between studies for mortality.

In assessing the robustness of results to the decision to include studies with high risk of bias, studies presenting with only low or moderate risk of bias were analysed. In so doing, a summary frequency of 16.9% was calculated from 13 included studies. This pooled proportion differs by nearly half when comparing to 28.3% and are thusly not robust, suggesting our results were influenced by risk of bias.

Regarding studies where maternal diagnosis of Chagas disease differed from WHO guidelines, a summary frequency of 31.2% was calculated from 28 included studies. This pooled proportion is similar to our primary findings and therefore results are robust and likely not influenced by method of maternal diagnosis for Chagas disease.

For infants diagnosed using the gold standard method, the estimated pooled proportion of infants symptomatic for cCD to all cCD infected infants was 18.7% versus 32.5% for infants diagnosed by an alternative method. Fewer infants were found symptomatic when diagnosed by the gold standard method, suggesting results may be influenced by infant diagnostic method. Additionally, the I^2^ values within each subgroup are considerably heterogenous (gold standard = 86.8% and alternative method = 78.9%), indicating substantial variability attributable to differences among diagnostic tests used for congenitally infected infants. Although overall highly heterogenous, it is interesting to note that both subgroups have a lower I^2^ value compared to the primary I^2^ value of 88.6%.

In the next subgroup analysis, the I^2^ statistic was calculated to measure the proportion of total variability attributable to heterogeneity in geographic area within each study. Based on geographic region, the pooled proportion of infants symptomatic for cCD to all cCD infected infants was 20.0% in studies based in Europe compared to 29.4% for studies based in Central/South America. Furthermore, heterogeneity was unlikely a factor with studies based in Europe (I^2^ = 10.2%); however, studies based in the Latin American and Caribbean region were considerably heterogenous (I^2^ = 91.3%). These results draw mixed conclusions regarding variability attributable to differences between geographic locations.

The proportion of infants symptomatic for cCD by their specific symptom to determine the individual symptom occurrence was calculated, with the top five most frequently occurring symptoms including hepatosplenomegaly, preterm birth, low birth weight, anemia, and jaundice. These symptoms are consistent with those reported in the literature [6-8, 10, 11, 79]. Considering heterogeneity, the I^2^ values ranged from moderately to considerably heterogenous (jaundice = 59.9%, low birth weight = 60.7%, preterm birth = 61.2%, anemia = 64.1%, and hepatosplenomegaly = 85.8%), indicating some variability attributable to differences in type of symptom displayed in the infant.
