## Supplemental Table 1 for "Estimation of the morbidity and mortality of congenital Chagas disease: a systematic review and meta-analysis"

**S1 Table.** Congenital cases morbidity characteristics

| **Study** | **Number Infected** | **Symptomatic**  n(%) | **Morbidity**  n(%) | | | | | |
| --- | --- | --- | --- | --- | --- | --- | --- | --- |
|  |  |  | **Hepato-splenomegaly** | **PTB** | **LBW** | **Anemia** | **Jaundice** | **Other** |
| Apt 2013 | 6 | 3(50.0) | 0(0.0) | 1(17.0)* | 0(0.0) | 0(0.0) | 0(0.0) | 1(17.0) - Transient hypotonia  2(33.0) - Respiratory symptomatology cyanosis and grunting  1(17.0) - Bronchopneumonia  1(17.0) Respiratory distress |
| Azogue 1991 | 78 | 21(27.0) | 35(45.0) | 0(0.0) | 0(0.0) | 0(0.0) | 11(14.0) | 4(5.0) - Edema  2(3.0) - Respiratory difficulties  4(5.0) - Other symptoms, undefined |
| Barona-Vilar 2012 | 8 | 1(13.0) | 1(13.0) | 0(0.0) | 0(0.0) | 0(0.0) | 0(0.0) | Not specified |
| Barousse 1978 | 1 | 1(100.0) | 1(100.0) | 1(100.0) | 1(100.0) | 0(0.0) | 0(0.0) | Not specified |
| Basile 2019^2^ | 28 | 4(14.0) | 0(0.0) | 0(0.0) | 0(0.0) | 0(0.0) | 3(11.0) | Not specified |
| Bern 2009 | 10 | 3(30.0) | 3(30.0) | 0(0.0) | 0(0.0) | 0(0.0) | 0(0.0) | Not specified |
| Bittencourt 1985 | 3 | 2(67.0) | 1(33.0) | 2(67.0) | 2(67.0) | 1(33.0) | 1(33.0) | 1(33.0) - Hyporeflexia  1(33.0) - Dyspnea  1(33.0) - Association with perinatal hemolytic disease |
| Buekens 2018 | 11 | 4(36.0) | 1(9.0) | 1(9.0) | 0(0.0) | 0(0.0) | 1(9.0) | 1(9.0) - Sepsis  1(9.0) Apgar <7 at 1-minute  1(9.0) - NICU admission  3(27.0) - PROM |
| Cardoso 2012 | 15 | 1(7.0) | 0(0.0) | 1(7.0) | 0(0.0) | 0(0.0) | 0(0.0) | Not specified |
| Castillo 1984 | 31 | 2(6.0) | 0(0.0) | 0(0.0) | 2(6.0) | 0(0.0) | 0(0.0) | Not specified |
| Flores-Chavez 2011 | 4 | 1(25.0) | 0(0.0) | 0(0.0) | 0(0.0) | 0(0.0) | 0(0.0) | 1(25.0) - Downs syndrome  1(25.0) - Congenital cardiopathy |
| Francisco-Gonzalez 2019 | 3 | 1(33.0) | 0(0.0) | 0(0.0) | 0(0.0) | 1(33.0) | 0(0.0) | 1(33.0) - Hydrops fetalis  1(33.0) - Ascites  1(33.0) Hemodynamic instability  1(33.0) - NICU admission |
| Freiliji 1995 | 71 | 25(35.0) | 13(18.0) | 0(0.0) | 0(0.0) | 1(1.0) | 0(0.0) | 5(7.0) - Sepsis  3(4.0) - Hepatitis  1(1.0) - Edema  3(4.0) - HIV co-infection  1(1.0) - Respiratory distress |
| Gimenez 2010 | 3 | 1(33.0) | 0(0.0) | 0(0.0) | 0(0.0) | 0(0.0) | 0(0.0) | 1(33.0) - Dilated cardiomyopathy  1(33.0) - Neuroblastoma |
| Martinez de Tejada 2009 | 2 | 1(50.0) | 1(50.0) | 0(0.0) | 0(0.0) | 0(0.0) | 0(0.0) | Not specified |
| Mayer 2010 | 18 | 9(50.0) | 7(39.0) | 0(0.0) | 0(0.0) | 0(0.0) | 0(0.0) | 4(22.0) - Cardiomyopathy |
| Messenger 2017¹ | 38 | 11(29.0) | 0(0.0) | 6(19.0) | 7(22.0) | 0(0.0) | 0(0.0) | 1 - Apgar <7 at 1-minute  5/32 - NICU admission  4(13.0) - PROM |
| Munoz 1982 | 2 | 2(100.0) | 0(0.0) | 1(50.0) | 1(50.0) | 1(50.0) | 0(0.0) | 1(50.0) - Central nervous system problems  1(50.0) - Possible overload of left ventricle |
| Nisida 1999 | 4 | 4(100.0) | 1(25.0) | 1(25.0) | 1(25.0) | 2(50.0) | 0(0.0) | 1(25.0) - Femur metaphysitis  1(25.0) - Seizure  1(25.0) - Stillbirth >20 weeks  1(25.0) - Spontaneous abortion <20 weeks |
| Otero 2012 | 1 | 1(100.0) | 1(100.0) | 1(100.0) | 1(100.0) | 0(0.0) | 0(0.0) | 1(100.0) - Cholestasis  1(100.0) - Cytolysis |
| Rodari 2018 | 1 | 1(100.0) | 0(0.0) | 1(100.0) | 0(0.0) | 1(100.0) | 0(0.0) | Not specified |
| Rubio 1962 | 1 | 1(100.0) | 1(100.0) | 0(0.0) | 0(0.0) | 0(0.0) | 0(0.0) | 1(100.0) - Broncopneumonia  1(100.0) - Hepatic steatosis |
| Salas 2007 | 58 | 15(31.0) | 0(0.0) | 0(0.0) | 0(0.0) | 13(22.0) | 0(0.0) | Not specified |
| Sosa-Estani 2009 | 8 | 2(25.0) | 0(0.0) | 0(0.0) | 1(13.0) | 0(0.0) | 0(0.0) | 1(13.0) - Dehydration  1(13.0) - Sepsis  1(13.0) Respiratory distress  1(13.0) - Gastroenteritis  1(13.0) - Scarce clear meconial liquid |
| Streiger 1995 | 9 | 6(67.0) | 3(33.0) | 5(56.0) | 6(67.0) | 0(0.0) | 1(11.0) | 1(11.0) - Respiratory difficulty  1(11.0) - Malnourished  1(11.0) - Neumopatia (lung disease) |
| Torrico 2004 | 71 | 36(51.0) | 21(30.0) | 3(4.0) | 4(6.0) | 0(0.0) | 0(0.0) | 9(13.0) - Cardiomegaly  2(3.0) - Apgar score 0 at 5-minute  8(11.0) - Neurological signs  6(8.0) - Anascara  6(8.0) - Petechia |
| Valenzuela 1984 | 11 | 4(36.0) | 0(0.0) | 0(0.0) | 3(27.0)* | 0(0.0) | 0(0.0) | Not specified |
| Valperga 1992 | 4 | 3(75.0) | 1(25.0) | 1(25.0) | 1(25.0) | 0(0.0) | 2(50.0) | 1(25.0) - Maxilofacial malformation  2(50.0) - SGA |
| Villablanca 1984 | 61 | 25(41.0) | 0(0.0) | 0(0.0) | 25(41.0) | 0(0.0) | 0(0.0) | Not specified |
| Zaidenberg 1993 | 12 | 12(100.0) | 12(100.0) | 5(42.0) | 0(0.0) | 10(83.0) | 10(83.0) | 2(17.0) - SGA |

*A value of 0 was inferred from missing data, as most studies only explicitly outlined symptoms that were present.*

*PTB counts any report of PTB <37 weeks), MPTB(32-36 weeks), VPTB(28-31 weeks), EPTB(<28 weeks)*

*LBW counts any report of LBW (<2500g), VLBW (<1500g), ELBW(<1000g)*

*Hepatosplenomegaly counts any report of hepatomegaly, splenomegaly, and hepatosplenomegaly*

*RDS = Respiratory distress syndrome*

*¹Study reported 38 infected cases and symptoms were reported out of 32 infected cases*

*^2^Unable to determine total number of infants with hepatosplenomegaly*

**Undefined LBW and PTB.*
