## Supplemental Table 2 for "Estimation of the morbidity and mortality of congenital Chagas disease: a systematic review and meta-analysis"

**S2 Table.** Congenital cases mortality characteristics

| Study | **#** | **Mortality** | |
| --- | --- | --- | --- |
|  |  | **Time** | **Cause** |
| Bittencourt 1985 | 1 | Birth | Stillbirth |
| Flores-Chavez 2011 | 1 | 9 months | Born with Down’s syndrome and congenital cardiopathy. Suddenly died after recovering from *T. cruzi* infection |
| Freiliji 1995 | 2 | 8 months | Respiratory distress |
|  |  | 14 months | Severe neurological damage |
| Nisida 1999 | 2 | Birth | Chagas disease |
|  |  | 30 Days | Chagas disease |
| Salas 2007 | 2 | Date undetermined | Not specified |
| Sosa-Estani 2009 | 2 | 4 months | Gastroenteritis and dehydration |
|  |  | 5 months | Pneumonia |
| Streiger 1995 | 1 | 2 months | Secondary septicemia and pneumococcal meningitis |
| Torrico 2004 | 4 | 24-48 hours | Chagas disease |
